# Cardiorespiratory fitness interventions for individuals with lower limb amputation: a scoping review

**DOI:** 10.1101/2022.12.22.22283838

**Authors:** K van Kammen, B.L. Seves, A.H. Vrieling, R. Dekker, A. van Dijk, P.U. Dijkstra, J.H.B. Geertzen

## Abstract

**Purpose:** This scoping review aimed to provide an overview of different cardiorespiratory training methods and their effects on cardiorespiratory fitness in persons with a lower limb amputation (LLA).

**Methods:** Studies were searched in PubMed, EMBASE, CINAHL, Cochrane Library and Web of Science. The search strategy comprised four search strings with terms related to ‘amputation’ or ‘limb loss’, ‘lower extremity’, ‘training’ or ‘exercise’, and ‘endurance’ or ‘aerobic’. Studies were included if they reported on persons with a LLA who followed a cardiorespiratory training. The Effective Public Health Practice Project tool was used for quality assessment.

**Results:** Nine studies (88 participants) were included, with weak (7 studies) to moderate (2 studies) quality. Duration of the programmes ranged from 1.5 to 26 weeks, with a frequency of 3-5 sessions per week. The intensity of the workload increased during the programmes in two studies. After training, maximal oxygen uptake (VO_2_max) had increased in all studies, in three studies significantly. The training effects were inconclusive for resting heart rate, peak heart rate, and blood pressure

**Conclusions:** Cardiorespiratory training methods – with tailored duration and intensity – tended to increase VO_2_max and are feasible in persons with a LLA but quality of the studies was weak.

**Implications for rehabilitation:**

1. VO_2_max of persons with a lower limb amputation can improve after cardiorespiratory training.
2. Training methods to improve cardiorespiratory fitness in persons with a lower limb amputation, in which training intensity is tailored to the patient (based on heart rate) are feasible.
3. Due to the low methodological quality of the included studies, this scoping review cannot provide an evidence-based overview of training methods to improve cardiorespiratory fitness in persons with a lower limb amputation.

## Introduction

Persons with a lower limb amputation (LLA) experience a decline in physical fitness due to decreased levels of physical activity after the amputation [1-5], and due to comorbidities, such as vascular disease and diabetes mellitus [6,7]. This decline in physical fitness is also related to a decrease in physical activity as a result of higher energy costs in walking with a prosthesis compared to walking with two sound legs [1,6,8]. Consequently, improving physical fitness by exercise training is expected to improve the ability to walk with a prosthesis in persons with a LLA [9,10]. According to the American College of Sport Medicine physical fitness consists of five components, namely: 1. cardiorespiratory fitness, 2. muscle strength, 3. muscular endurance, 4. flexibility and 5. body composition [11]. Since the burden on the cardiorespiratory system of persons with a LLA is high, especially in persons with an amputation due to vascular disease [1], the current scoping review only focuses on cardiorespiratory fitness (usually expressed in maximal oxygen uptake [VO_2_max], heart rate and blood pressure) in persons with a LLA. Also, the improvement of cardiorespiratory fitness seems to be a key factor related to the functional outcomes of persons with a LLA, such as walking ability with a prosthesis [3,12,13].

Worldwide amputations in persons with a LLA is mostly due to vascular deficiency, ranging from 25% to 90% [7,14]. Training methods to improve cardiorespiratory fitness in persons with a dysvascular LLA might be different compared to persons with a traumatic LLA since generally the vascular problems have affected other organs as well in persons with a dysvascular amputation [15]. Also, persons with a dysvascular LLA are on average older compared to persons with a traumatic LLA [1].

In the Netherlands, the objective assessment of cardiorespiratory fitness and use of exercise testing is not routine among persons with a LLA in clinical rehabilitation. A detailed description of training methods to improve cardiorespiratory fitness is limited in the Dutch guideline for persons with a LLA [16]. Additionally, literature on specific training methods for dysvascular LLA and for traumatic LLA is scarce [17]. Therefore, an evidence-based overview of training methods to improve cardiorespiratory fitness in persons with a LLA is needed to recommend clinicians on this topic.

Rehabilitation professionals may use such an overview to make an evidence-based decision on training methods aiming to improve cardiorespiratory fitness in persons with a LLA. Furthermore, such an overview will identify knowledge gaps on cardiorespiratory training methods in persons with a LLA. Therefore, the aim of this scoping review is to provide a detailed overview of different cardiorespiratory training methods and its effects on the cardiorespiratory fitness in persons with a LLA.

## Methods

This manuscript is reported according to the PRISMA guidelines for scoping reviews [18]. Prior to the review a protocol was specified (online supplemental file 1).

### Searches

Studies were searched in PubMed, EMBASE, CINAHL, Cochrane Library and Web of Science. The search strategy was composed of four search strings; the first was related to the terms “amputation” and “limb loss”, the second to “lower extremity”, the third to “training” and “exercise” and the last string was dedicated to “endurance” and “aerobic”. The search strings were combined using “AND”. The search terms included MeSH terms and free text. The search strategies were generated together with an information specialist (librarian) with advanced experience in developing search strategies for systematic reviews. No time or language restrictions were applied. The final search was performed on May 10^th^, 2021 (online supplemental file 1).

### Eligibility criteria

Full text studies were included if they reported on persons with a LLA who followed a training program to improve the cardiorespiratory fitness, and if amputation level was proximal to Syme level. Books, notes, letter to editors, conference abstracts and (systematic) reviews were excluded. Reference lists of systematic reviews were checked prior to exclusion to include relevant studies that were missed in the database search.

### Study selection

The study selection was pilot tested by two reviewers before each step of the review. The pilot focused on the interpretation of the inclusion criteria that are defined in the protocol (online supplemental file 1). Two reviewers (AvD & JHBG) assessed titles and abstracts independently. Titles and abstracts were excluded if both reviewers rejected them. In case of disagreement, reviewers discussed with a third independent reviewer (PUD) until consensus was reached. After the selection based on titles and abstracts it was determined if papers required translation. Subsequently, two reviewers assessed the full text (AvD & AHV), and articles were excluded if they were rejected by both reviewers. In case of disagreement, a third independent reviewer (PUD) performed full text assessment of the article. The reference lists of included studies were checked if relevant studies were missed in the database searches and were assessed on the same way. Cohen’s kappa (k) was calculated as a measure for the inter-rater reliability.

### Risk of bias (quality) assessment

The quality assessment was independently performed by two reviewers (AvD & RD) using the Effective Public Health Practice Project (EPHPP) tool [19]. The tool was pilot tested on excluded studies.

### Primary outcome(s) and data extraction

As the current scoping review aimed to acquire insight into (1) types of interventions to improve the cardiorespiratory fitness in persons with a LLA, and (2) the effects of the interventions on cardiorespiratory fitness, data extracted in terms of reported outcomes were resting heart rate, peak heart rate, VO_2_max and blood pressure. To determine facilitators, barriers, safety and feasibility of the interventions, data on the setting of the intervention was extracted (online supplemental file 1).

Data was extracted by four reviewers (AvD, RD, KvK, BS) independently using a form developed for this study. The data extraction form was pilot tested by two reviewers (AvD & RD). In case of missing data or incomplete studies, the first reviewer (AvD) contacted the corresponding authors by email.

## RESULTS

### Selected studies

After duplicate removal, 1122 references were assessed for eligibility. A total of 41 studies were selected for the full-text assessment (Cohen’s kappa = 0.782) (figure 1). Nine studies (table 1) were included (Cohen’s kappa = 0.407) [4,20-27], of which one article required translation from French to English [26].

**Table 1.** Characteristics of the included studies

| Authors<br>country | (year) | Study design | n(%male) | Drop out | Age<br>mean (SD) [range] | Cause | TF | TT | F | TF+TT | Uni/Bi |
| --- | --- | --- | --- | --- | --- | --- | --- | --- | --- | --- | --- |
| James | (1973) | Pre-post | 11(100%) | NA | 37.5(9.0) | NA | 11 |  |  |  | 11/0 |
| Sweden |  | +Control group | 11 | NA | 44 [23-59] | NA |  |  |  |  |  |
| Miller | (1976) | Pre-post | 7(57%) | NA | 65.9(9.1) |  | 3 | 4 |  |  | 7/0 |
| USA |  |  |  |  |  |  |  |  |  |  |  |
| Adler | (1987) | Case report | 1(100%) | NA | 63 | V |  | 1 |  |  | 0/1 |
| USA |  |  |  |  |  |  |  |  |  |  |  |
| Bostom | (1987) | Case report | 1 (100%) | NA | 73 | V | 1 |  |  |  | 1/0 |
| USA |  |  |  |  |  |  |  |  |  |  |  |
| Pitetti | (1987) | Pre-post | 10* | 10 | 39.3(8.1) | T | 3 | 4 | 1 | 2 | 8/2 |
| USA |  | +Control group | 5 | NA | 42 |  | 2 | 4 | 1 |  | 7/0 |
| Davidoff | (1992) | Pre-post | 25(100%) | NA | 63.4(8.4) | V | 7 | 18 |  |  | 25/0 |
| USA |  |  |  |  |  |  |  |  |  |  |  |
| Paré | (1996) | Pre-post | 18(100%) | 5 | 64.0(11.7) | V | 7 | 10 | 1 |  | 18/0 |
| Canada |  |  |  |  |  |  |  |  |  |  |  |
| Chin | (2001) | Pre-post | 14 | NA | 39.8(12.4)# | T | 14 |  |  |  | 14/0 |
| Japan |  | +Control group | 10 | NA | 41.2(18.4)# | T | 10 |  |  |  | 10/0 |
| Donachy | (2004) | Case report | 1(100%) |  | 40 | T |  | 1## |  |  | 1/0 |
| USA |  |  |  |  |  |  |  |  |  |  |  |

Number (n), not available (NA), vascular (V), trauma (T), transfemoral (TF), transtibial (TT), foot (F), unilateral (Uni), bilateral (Bi), \* 20 persons started the program, persons who completed the training program were reported. #: Reported as standard error by Chin et al 2001 [4], ##: Also a shoulder disarticulation

**Figure 1.**
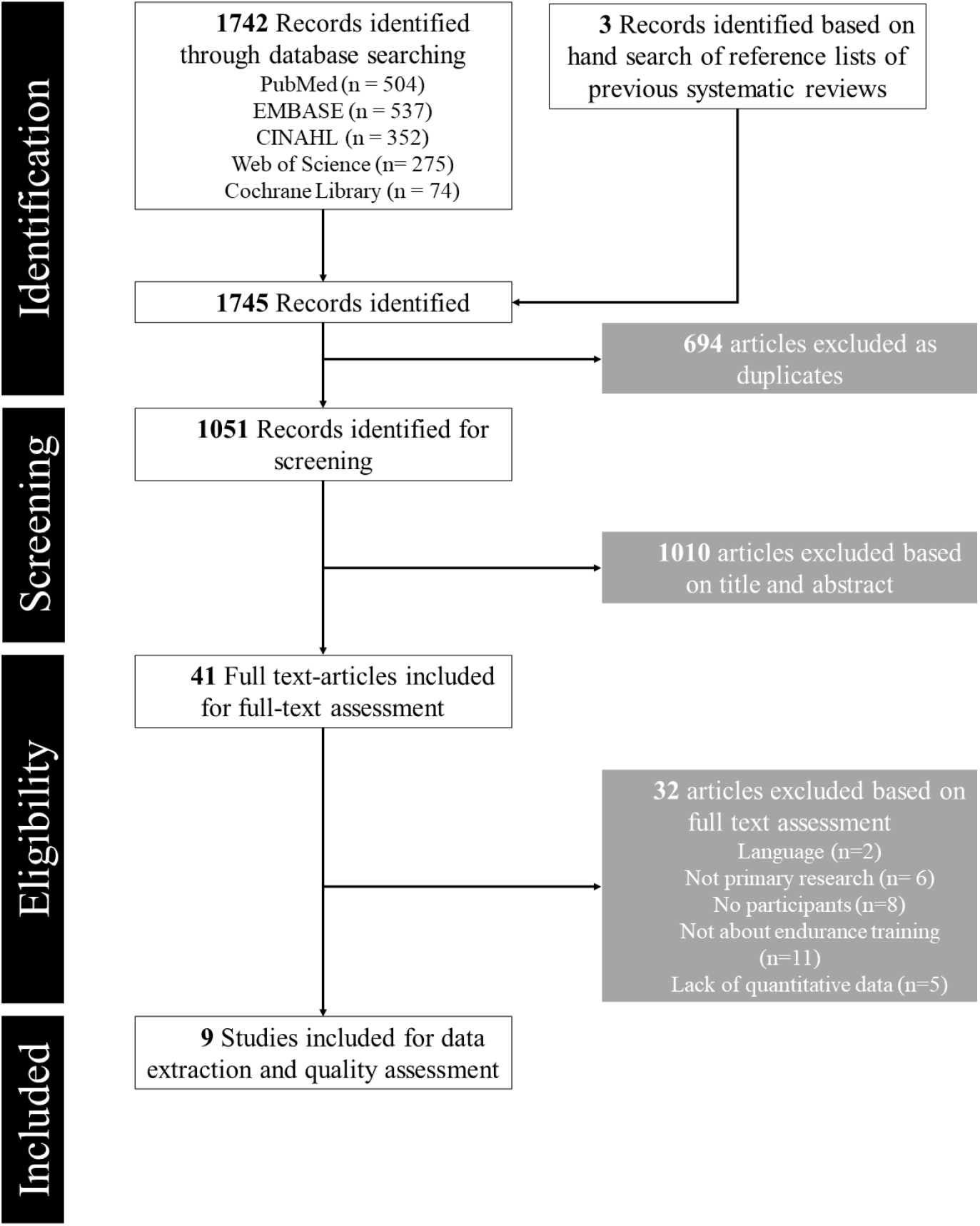
Prisma flow chart (n=number of studies)

Studies were performed between 1973 and 2004 and included three case studies and six clinical evaluations of which three include a control group. The quality of seven studies was rated weak (table 2). Two clinical evaluations, one without control group [23] and one with control group [4] were rated as moderate. In seven studies training was conducted in a clinical setting. In the other two studies training was conducted at home, with guidance of a therapist once or twice a week by phone.

**Table 2.** Quality of the included studies, based on the Effective Public Health Practice Project Tool [19]

| Author, year | Selection bias | Study design | Confounders | Blinding data | Collection | Drop-outs | Global Rating |
| --- | --- | --- | --- | --- | --- | --- | --- |
| James, 1973 | w | m | w | w | w | m | w |
| Miller, 1976 | m | m | w | m | w | s | w |
| Adler, 1987 | m | w | w | m | s | m | w |
| Bostom,1987 | m | m | w | m | s | s | m |
| Pitetti, 1987 | w | m | w | w | s | w | w |
| Davidoff, 1992 | m | m | w | m | s | w | w |
| Pare, 1996 | w | m | w | w | w | m | w |
| Chin, 2001 | w | m | s | m | s | s | m |
| Donachy, 2004 | w | w | w | m | s | m | w |
| Weak (w), moderate (m), strong (s) |  |  |  |  |  |  |  |

### Participants

In the studies, a total of 103 participants started training of which fifteen dropped out, leading to a total research population of 88 participants (mean age ranged from 37.5 to 73.0) (table 1). In a clinical study the reason for dropouts (n=5) was inability to reassess subjects for technical reasons [26]. In a home-based study participants (n=10) did not adhere to the training program due to personal and motivational reasons [24]. Six studies included males only, and in two studies females participated also. Two studies did not report gender of the participants. Sample size ranged from 1 to 25, and six studies were performed in the USA. Reason for amputation was vascular disease in four studies and trauma in three studies. Two studies did not report reason for amputation. In seven studies the participants had a unilateral LLA.

### Physical exercise assessment

In all studies a pre- and post-exercise-test was performed to measure the changes in physical fitness of the participants. The used ergometers were a bicycle ergometer, Aire-Dyne ergometer, arm ergometer and a treadmill (table 3). Three studies used multiple ergometers for the tests. All tests were maximal tests. In all studies, participants were monitored with an ECG monitor and blood pressure measures during testing, but the outcomes of these measurements were not always reported.

**Table 3.**
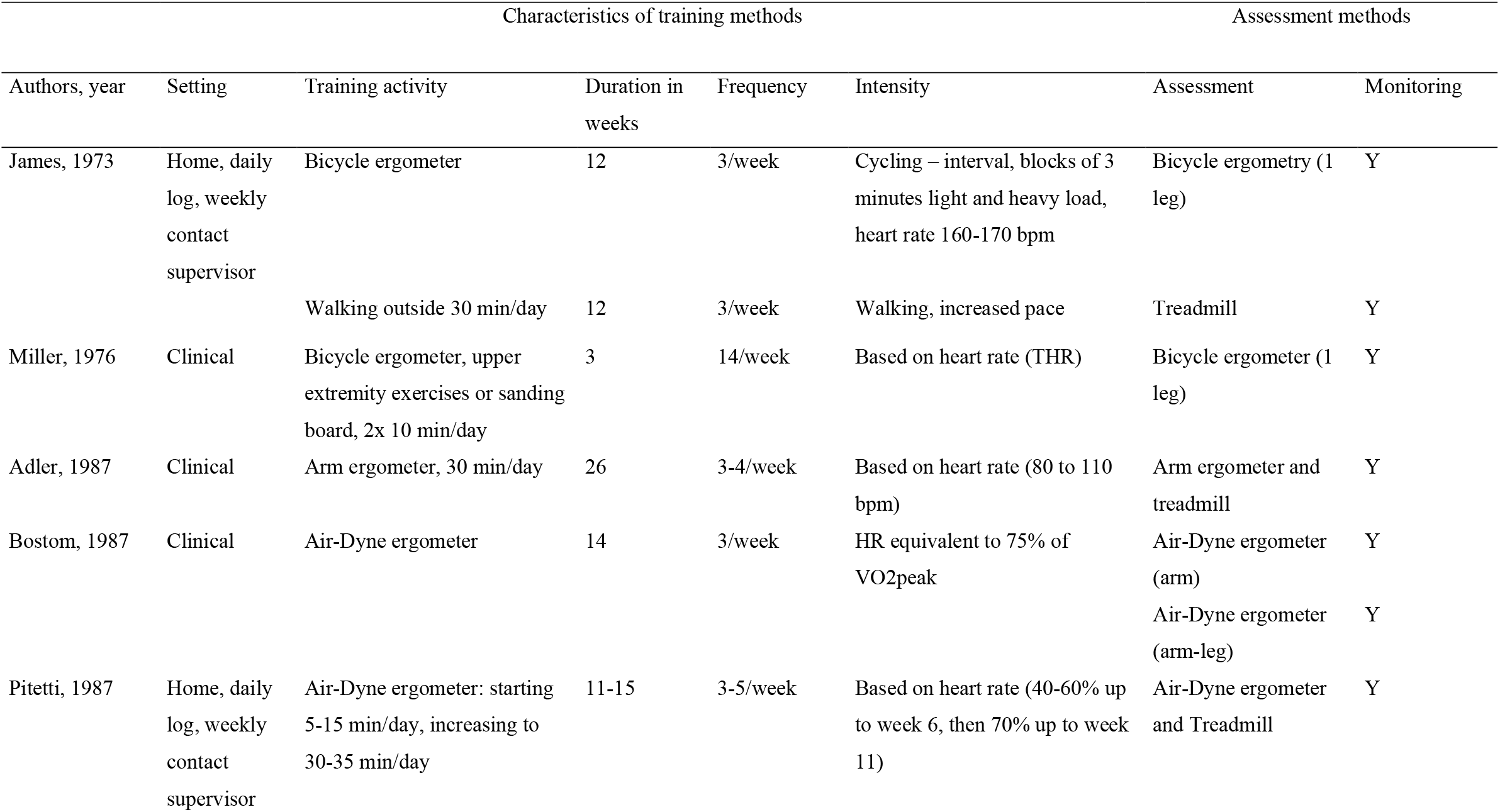
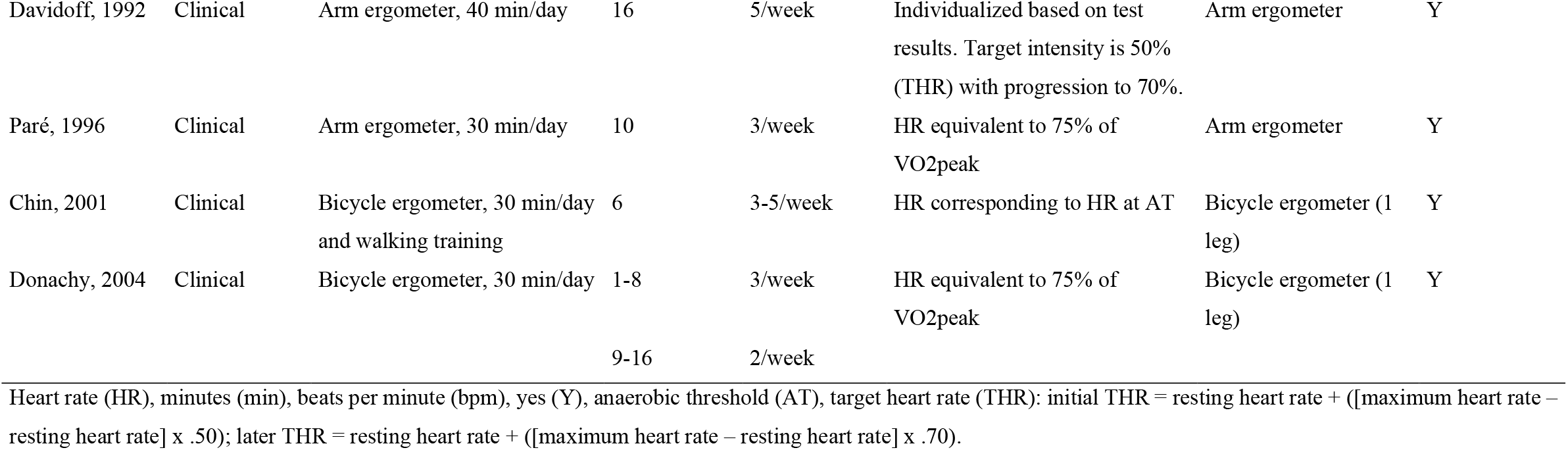
Characteristics of training methods and assessment methods in the included studies.

### Intervention program

The intensity of the workload increased during the programme in two studies. All studies provided an individual tailored training program based on heart rate during exercise testing. The duration of the programs ranged between 3 to 26 weeks with 3 to 14 sessions per week. The duration per session ranged from 5 to 40 minutes. The training activities were performed on bicycle ergometer (4 studies), arm ergometer (3 studies) and Aire-Dyne ergometer (2 studies).

### Primary outcomes

#### Resting heart rate

A total of four studies reported pre- and post-measurements of resting heart rate (table 4). These studies showed that the resting heart rate remained stable [20] or decreased [21,24,25] after the intervention. In one of those studies the decrease was statistically significant [24].

**Table 4.** Outcome measures in the included studies

| Author,<br>year | Resting heart rate mean<br>(SD) bpm |  |  | Peak heart rate<br>mean (SD) bpm |  |  | VO<sub>2</sub>max<br>mean (SD) ml/kg/min |  |  | Blood pressure diastolic / systolic<br>mean (SD) mmHG |  |  |
| --- | --- | --- | --- | --- | --- | --- | --- | --- | --- | --- | --- | --- |
|  | Pre | Post | %change | Pre | Post | %change | Pre | Post | %change | Pre | Post | %change |
| James,<br>1973 | 85<br>(2) | 85<br>(10.8) | 0%<sup>s</sup> | 160<br>(18.1) | 173<br>(16.3) | +8.1%<sup>s</sup> | 24.3<br>(5.2) | 32.0<br>(5) | +31.7%<sup>s</sup> |  |  |  |
| Miller,<br>1976 | 85.7 | 75 | -12.5%<sup>s</sup> |  |  |  |  |  |  |  |  |  |
| Adler,<br>1987 |  |  |  |  |  |  | 5.5 | 15.8 | +187.3%<sup>s</sup> | Rest: 70 / 140 | Rest: 80 / 120 | Rest: +14.3%<sup>s</sup> / -14.3%<sup>s</sup> |
| Bostom,<br>1987 |  |  |  | 155<br>160 | 145<br>155 | -6.5%<sup>s</sup><br>-3.1%<sup>s</sup> | 14.9<br>15.3 | 14.0<br>19.1 | -6.0%<sup>s</sup><br>+24.8%<sup>s</sup> | Peak: 110/210<br>Peak: 96/194 | Peak: 90/210<br>Peak: 90/194 | Peak: -18.2%<sup>s</sup> / -7.6%<sup>s</sup><br>Peak: -6.3%<sup>s</sup> / +8.3%<sup>s</sup> |
| Pitetti,<br>1987 | 81<br>(5) | 70<br>(5) | -<br>13.6%* | 154<br>(7) | 170<br>(6) | +10.4%* | 23.1 | 29.4 | +27.3%*** | Rest: 85 (3) / 135 (3) | Rest: 82 (3) / 131 (6) | Rest: -3.5% n.s. / -3.0% n.s. |
| Davidoff,<br>1992 | 87<br>(13) | 83<br>(13) | -4.6%<br>n.s. | 119<br>(13) | 117<br>(16) | -1.7%<br>n.s. |  |  |  | Rest: 75.8 (10.3) / 122 (19)<br>Peak: 81.6 (14) / 152 (29) | Rest: 76.9 (9.6) / 129 (18)<br>Peak: 84 (13) / 163 (21) | Rest: +1.5% n.s. / +5.7% n.s.<br>Peak: +2.9% n.s. / +7.2%* |
| Paré,<br>1996 |  |  |  | 134<br>(11) | 129<br>(8) | -4%** | 18.7<br>(0.8) | 22.2<br>(1.3) | +19%** |  |  |  |
| Chin,<br>2001 |  |  |  |  |  |  |  |  | +36.5%* |  |  |  |
| Donachy,<br>2004 |  |  |  |  |  |  | 29.73 | 38.73 | +30.3%<sup>s</sup> |  |  |  |

Standard deviation (SD), beats per minute (bpm), maximal oxygen uptake (VO<sub>2</sub>max), millimetre of mercury (mmHG)
\*=p<.05, \*\*=p<.01, \*\*\*=p<.001, n.s.=not significant, <sup>s</sup>=descriptive statistics only

#### Peak heart rate

Five studies reported pre- and post-measurements of peak heart rate. Three studies showed a decrease of peak heart rate [23,25,26]. In one of those studies the decrease was statistically significant [26]. Two studies reported an increased peak heart rate after the training [20,24]. In one of those studies, results were statistically significant [24].

#### VO_2_max

In seven studies, VO_2_max was assessed and reported pre- and post-training measurements and/or percentage change between pre- and post-training measurements [4,20,22-24,26,27], all showed an increase in VO_2_max which was statistically significant in three studies [4,24,26].

#### Blood pressure

Pre- and post-training assessment of blood pressure was reported in four studies. An increase in diastolic blood pressure in resting conditions was reported in two studies [22,25], and one study reported a decrease [24]. An increase in systolic blood pressure in resting conditions, was reported in one study [25] and two reported a decrease [22,24]. None of the results were significant. An increase in peak blood pressure of both diastolic and systolic blood pressure was found in one study [25] and another found decrease in both diastolic and systolic blood pressure [23]. In one study the increase in systolic blood pressure was statistically significant [25].

## DISCUSSION

This scoping review aimed to provide a detailed overview of different cardiorespiratory training methods and their effects on the cardiorespiratory fitness in persons with a LLA. Overall, the current scoping review found weak evidence to support that training methods to improve cardiorespiratory fitness increase cardiorespiratory fitness in persons with a LLA.

### Training effects

At the individual level the current scoping review found that the training methods did generally improve cardiorespiratory fitness in persons with a LLA. The VO_2_max increased after the intervention in all included studies that reported this measure, which was statistically significant in three studies [4,24,26]. However, for the remaining primary outcomes, the interpretation of the results is less straightforward. The resting heart rate decreased or stayed the same after the intervention, but a significant decrease was only found in the study of Pitetti et al. [24]. The peak heart rate significantly increased and significantly decreased after the intervention, respectively in the study of Pitetti et al. [24] and Paré et al. [26]. The effects of the intervention on blood pressure were inconclusive and non-significant, for both resting blood pressure as peak blood pressure. The above-mentioned contradictory, inconclusive or non-significant findings can be explained by the small sample size of the study groups and thus the limited statistical power, and by the large variation in training activity, duration, frequency, and intensity in the included studies.

From the results of this review, we cannot conclude whether a training effect is larger after a long training period, after training on high intensity or after training either on a bicycle ergometer, an Aire-Dyne ergometer or an arm ergometer, due to the limited number of studies and the low methodological quality. The large number of drop-outs (10 out of 20 participants in the intervention group) in the study of Pitetti et al. [24], might explain the large significant training effects found in that study.

It has been stated in previous literature that endurance training methods should be tailored to the reason of amputation: vascular or traumatic [15]. However, based on this scoping review, it is impossible to compare the effects of endurance training methods on cardiorespiratory fitness in persons with a dysvascular versus traumatic amputation, since training effects might be attributed to the cause of the amputation or to training duration, training frequency, training intensity and training activity between studies. To investigate if different cardiorespiratory training methods should be developed for persons with a dysvascular or traumatic amputation, we recommend future research to perform a before-after intervention design and to include two intervention groups, one with persons with a dysvascular amputation and one with persons with a traumatic amputation.

### Feasibility of the training methods

Despite the intensive training frequency (on average 30 min/day, 3-5 times per week) and duration (on average 12 weeks), the different training methods described in the included studies seem feasible since only 15 participants dropped-out of the 103 included participants. Remarkably most drop outs occurred in an home-based programme due to personal and motivational reasons [24]. These findings might implicate that the interventions in clinical settings facilitate adherence to the training program. This interpretation, however, should be considered with caution due to the small number of studies included in this scoping review.

In addition, the included studies did not report adverse effects or any safety issues during the training program, making it difficult to draw conclusions on the safety of the interventions. We recommend future research to report whether adverse events occur during cardiorespiratory training. Also, we like to stress that future research might determine facilitators and barriers of home-versus clinical based training programs to confirm feasibility and safety of the interventions.

### Limitations and strengths

The conclusions of this review are shaped by the included studies. The major strength of this scoping review is the thorough approach applied for the search and inclusion strategy. However, some limitations do need to be addressed. Firstly, the number of studies identified for inclusion was low and methodological quality low to moderate. Secondly, in most included studies the sample size was too low to support conclusions with statistics and in other included studies the performed statistical analyses were poor. Thirdly, the included studies were published in 2004 and earlier. This might be explained by the specific scope of this review on endurance training and its effects on cardiorespiratory fitness. As a result, studies that investigated the effects of cardiorespiratory fitness after a combined endurance and strength training program were excluded. Recent studies on isolated endurance programs are probably scarce since training principles in persons after LLA have shifted from endurance training only to combined endurance and strength training programs over the past years. In a recent systematic review (2021), a broader perspective of exercise training programmes and its effects on physical fitness and functionality in persons with a LLA was summarized and included more recent publications [17]. Fourthly, the generalisability of the findings to a broad population of persons with a LLA is poor, since most participants in the included studies were male and had a unilateral LLA. Lastly, most studies were conducted in the USA, and implications derived from the findings in these studies may not apply to clinical settings in other countries and social or cultural context.

### Clinical implications and recommendations for future research

We did find that VO_2_max of persons with a LLA can improve after cardiorespiratory training. However, this review cannot provide an evidence-based overview for clinicians of training methods to improve cardiorespiratory fitness in persons with a LLA, due to low methodological quality of the included studies. However, training methods to improve cardiorespiratory fitness in persons with a LLA, in which training intensity was tailored to the patient (based on heart rate) seem to be feasible. The generalization of the results to a broad population of persons with a LLA is limited, due to the predominant profile of males with a unilateral amputation in the included studies. Therefore, we recommend future research to include a more heterogeneous group of persons with a LLA (male/female, unilateral/bilateral, onset of amputation surgery, and comorbidities). Moreover, there is a need for studies with high methodological quality, including a control group and follow-up measurements after the intervention to investigate the confounding effect of a control group and whether effects sustain on the long-term. Besides, we recommend future research to transparently report program details (such as how the program was tailored or how exercise progression was determined), safety measures and/or facilitators/barriers of the program, so that clinicians can reproduce the intervention in clinical rehabilitation.

## Conclusion

The current scoping review found weak evidence to support that training methods to improve cardiorespiratory fitness increase cardiorespiratory fitness in persons with a LLA. Future research should include a more heterogeneous sample of persons with a LLA, an intervention group with a combined endurance and strength training program, a control group, a follow-up measurement after the intervention, and should have high methodological quality.

## Data Availability

All included and excluded articles in this scoping review are available upon request to the authors.

