## Supplementary material for "Cardiorespiratory fitness interventions for individuals with lower limb amputation: a scoping review": online supplemental file 1

### Scoping review protocol
**Version: 2019**

---

#### Review title:
Cardiorespiratory fitness interventions for individuals with lower limb amputation: a scoping review

---

**Department of Rehabilitation Medicine, University Medical Center Groningen, Groningen, The Netherlands**

##### Background
Individuals with a lower limb amputation experience a decline in physical fitness due to limited physical activity prior to and after the amputation, cardiovascular disease and smoking habits. Studies demonstrated that cardiorespiratory fitness in amputees was lower than in their able-bodied peers. (1-5) Cardiorespiratory fitness is described by the World Health Organization (2010) as *"the ability of the circulatory and respiratory systems to supply oxygen during sustained physical activity. Usually expressed as measured or estimated maximal oxygen uptake (VO2max)*".(2) A lower cardiorespiratory fitness causes problems in walking with a prosthesis because energy expenditure in walking with a prosthesis is much higher than in walking with two sound legs.(1,6) To walk with a prosthesis at a functional level of activity, it is very important that the amputee is able to meet the high energy expenditure demands. Ascending levels of amputation appear to be *associated* with increased energy expenditure in walking.(3)

The burden on the cardiorespiratory system of amputees is high, especially in patients with an amputation due to vascular disease.(1) If it is possible to improve the physical fitness of the amputee, a reduction of the burden on the cardiorespiratory system of the amputee can be expected. Therefore, improvement of physical fitness seems to be an important factor related to functional outcome of the amputee.(3,8)

The American College of Sports Medicine (ACSM) guidelines for individuals with chronic diseases and disabilities, including individuals with a lower limb amputation recommend that achieving an aerobic training effect requires exercising of large muscle groups at a frequency of 3-5 times a week, for a duration of 20-60 minutes at an intensity of 40-70% heart rate reserve.(7) However, there is relatively little known about how an actual training program should be composed for this patient group.

##### Relevance
There is no overview of training methods to improve the cardiorespiratory fitness of individuals with a lower limb amputation. Such an overview can be useful for healthcare providers to make an evidence based decision for a certain fitness improving intervention for patients with a lower limb amputation.

Besides that it is of great importance to identify the gaps in knowledge about training methods for individuals with a lower limb amputation.

#### Research question

*Main question*

1. What types of interventions to improve the cardiorespiratory fitness of lower limb amputees are addressed in the literature?

*Sub questions*

2. What are the key program components of these interventions?
3. In what settings are these interventions provided?
4. Are these interventions effective, and what is de scale of improvement of the cardiorespiratory fitness of lower limb amputees?
5. Which measures are used to assess the improvement of  the cardiorespiratory fitness of lower limb amputees?
6. What is the cause of the lower limb amputation?
7. What are the barriers and facilitators to effective implementation of these interventions?
8. Are these interventions safe?

#### PCC

P =(population) Lower limb amputees
C = (concept) Interventions to improve cardiorespiratory fitness
C = (context) Rehabilitation to improve physical fitness

#### Search strategy

The search strategy is developed for the search in PubMed and will be translated to the other databases mentioned below (appendix I)

| **Database 1 name: PubMed** |
|---|
| **Search strategy:**<br>("amputation"[MeSH] OR "Amputees"[Mesh] OR "artificial limbs"[MeSH] OR amput*[tiab] OR limb loss[tiab] OR leg prosthes*[tiab] OR artificial leg[tiab])<br>AND<br>("lower extremity"[MeSH] OR lower extremit*[tiab] OR lower limb*[tiab] OR leg[tiab] *OR legs[tiab] OR one-legged[tiab]* OR below the knee[tiab] OR below knee[tiab] OR transtibial[tiab] OR trans-tibial[tiab] OR transfemoral[tiab] OR trans-femoral[tiab] OR Syme[tiab] OR "Tibia"[Mesh] OR tibia*[tiab])<br>AND<br>("Exercise"[MeSH] OR "Physical Therapy Modalities"[MeSH] OR "Exercise Test"[Mesh] OR "Endurance Training"[Mesh] OR "Amputation/rehabilitation"[Mesh] OR "Amputees/rehabilitation"[Mesh] OR exercis*[tiab] OR training[tiab] OR prosthetic rehabilitation[tiab])<br>AND<br>("Physical Endurance"[Mesh] OR "Endurance Training"[Mesh] OR "Physical Fitness"[Mesh] OR "Heart Rate"[Mesh] OR "Oxygen Consumption"[Mesh] OR "Energy Metabolism"[Mesh] OR oxygen[tiab] OR Vo2[tiab] OR metabolic cost*[tiab] OR endurance*[tiab] OR stamina[tiab] OR cardio*[tiab] OR aerobic[tiab] OR cardiac[tiab] OR exertion*[tiab] OR physical performance[tiab] OR exhaustion*[tiab] OR heart rate[tiab] OR energy |

| |
|---|
| expenditure[tiab]) |
| **Search result (# items retrieved): 504**<br>**Search date: 10-05-2021** |

#### Databases

- PubMed
- EMBASE
- CINAHL
- Web of Science

The reference list of the most relevant studies will be checked on missing data. Grey literature will not be included in the screening

#### Selection criteria

| **Title – abstract screening** | | | | |
|---|---|---|---|---|
| **Assessor** | **Inclusion criteria number** | Yes | No | Unclear |
| **Patient/participants:**<br>Do the participants have a lower limb amputation? (Syme level and more proximal) | **1** | | Only **no** when population specified with no lower limb amputation or distal to Syme level | |
| **Intervention:**<br>Is there a training program to improve cardiorespiratory fitness mentioned? | **2** | | Only **no** when training program or related terms (e.g. work-out, exercise, training, fitness, rehabilitation) are not mentioned | |
| **Method:**<br>Is the cardiorespiratory fitness of persons with LLA trained/Improved? | **3** | | Only **no** when PA or related terms (e.g. cardiorespiratory, Vo2Max, energy expenditure, endurance, stamina ) are not mentioned | |
| | **Exclusion criteria number** | Yes | No | Unclear |
| **Patients/participants:**<br>Is the study done in anything other than humans? | **1** | Only **Yes** when explicitly stated animal study or computer model without | | |

| | | |
|---|---|---|
| | | participants |
| **Study type:**<br>Is it a systematic review | **2** | If yes, then only review the reference list |

**Rules for going to the full-text phase:**

*Inclusion criteria*

All three Yes – full-text phase

Two Yes- full-text phase

One yes - exit

Unclear – full-text phase

*Exclusion criteria*

one yes – exit (systematic reviews will be only be reviewed for their reference list)

two yes – exit

| **Full text screening** | | | | |
|---|---|---|---|---|
| **Assessor** | **Inclusion criteria number** | Yes | No | Unclear |
| **Patient/participants:**<br>Do the participants have a lower limb amputation? (Syme level and more proximal) | **1** | | Only **no** when population specified with no lower limb amputation or distal to Syme level | |
| Where patients/participants aged ≥ 18 years?* | **2** | | Only **no** when term children or age range <18 | |
| **Intervention:**<br>Is there a training program to train cardiorespiratory fitness mentioned? | **3** | | Only **no** when training program or related terms (e.g. work-out, exercise, training, fitness, rehabilitation) are not mentioned | |
| **Method:**<br>Is the cardiorespiratory fitness of persons with LLA trained/Improved? | **4** | | Only **no** when PA or related terms (e.g. cardiorespiratory, Vo2Max, energy expenditure, endurance, stamina ) are not mentioned | |
| | **Exclusion** | Yes | No | Unclear |

| | **criteria number** | |
|---|---|---|
| **Patients/participants:**<br>Is the study done in anything other than humans? | **1** | Only **Yes** when explicitly stated animal study or computer model without participants |
| Do the patients/participants have an intellectual, sensory, cognitive or mental disability | **2** | Only **Yes** when explicitly stated that patients have such a disability |
| **Method:**<br>Is the cardiorespiratory fitness of persons with LLA measured as a functional or performance outcome? | **3** | Performance measured by a non-exercise test: e.g. 10 meter walking test, timed up and go<br>Functional e.g. gait analysis, range of motion |
| Are prosthetics compared in relation to cardiorespiratory fitness? | **4** | Only **Yes** when two prosthetic components are compared |

*Incase less than 20 articles to include, we will reconsider to include studies performed on children or age range <18

**Rules to accept or decline an article:**
*Inclusion criteria*
All four Yes – include in study
Unclear – discuss with other assessor
*Exclusion criteria*
one yes – exit

Language restriction: There are no language restrictions, after the abstract selection we will decide whether studies need to be translated.
Time restriction: No time restriction

---

#### Protocol for study selection

##### Title and abstract screening

Title and abstract of papers identified and those from reference check will be assessed by two assessors independently for inclusion criteria . Intra-rater agreement for title abstract screening will be calculated as % overall agreement and Cohen's Kappa (K: 0.00 – 0.20 = slight; 0.21 – 0.40 = fair; 0.41 – 0.60 = moderate; 0.61 – 0.80 = good; 0.81 – 1.00 = very good (8)). In case of disagreement assessors will discuss with a third arbiter to reach consensus. In case no consensus can be reached the article goes to the full text phase to be screened.

##### Full-text screening

Full text versions of the potentially eligible papers will be retrieved and independently assessed for by two assessors. Intra-rater agreement for full-text screening will be calculated as % overall agreement and Cohen's Kappa (K: 0.00 – 0.20 = slight; 0.21 – 0.40 = fair; 0.41 – 0.60 = moderate; 0.61 – 0.80 = good; 0.81 – 1.00 = very good (8)). The assessors will discuss studies in case of disagreement during a consensus meeting and decide to include or not to include. In case no consensus is reached, a third arbiter will be consulted to reach consensus.

In case of two or more papers describing the same study (same participants, measurements) one paper will be included . The most complete paper fitting  best the research question, with the largest sample size most recent? Longest follow-up?) will be included.

---

#### Quality assessment

The quality assessment will be performed with use of the EPHPP tool.

#### Data extraction

| **Data extraction framework** | | |
|---|---|---|
| *Main category* | *Subcategory* | *Description* |
| **Evidence source Details and Characteristics** | | |
| 1.Authors | | |
| 2.Title | | |
| 3.Journal | | |
| 4.Year of publication | | |
| 5.Objective(s) of the study | | Stated objectives of the study |
| 6.Type of study | | |
| 7. Country | | |
| **Inclusion & exclusion criteria** | | |
| 8.Population | Target population<br><br><br>Age<br><br>Gender<br><br>Cause of amputation<br><br>Level of amputation<br><br>Recruitment | Specify if the intervention targets individuals within subpopulation groups or the broad population<br>Specify the age groups covered by the study<br>Specify the gender groups covered by the study<br>Specify the cause of the amputation (eg, trauma, diabetes, cancer)<br>Specify the level of the amputation of the participants included in the study<br>Specify how the participants were |

| | | |
|---|---|---|
| | | recruited for the study and what were the completion rates |
| 9.Concept (intervention) | Type of intervention<br><br>Delivery of the intervention<br><br>Length of training, frequency, amount of weeks and intensity of the intervention | Description of intervention (type of exercise, dose, combined with other program,)<br><br>By whom and how was the intervention delivered<br><br>Description of the Length of training, frequency, amount of weeks and intensity of the intervention |
| 10.Context (setting) | | Describe the study setting (e.g. clinical setting, primary health care, home setting) |
| **Details/Results extracted from source of evidence** | | |
| 11.Reported outcomes | | Describe the intervention outcomes in the study (improved physical fitness, daily activity, mobility, improved vo2max) |
| 12. Types of assessment | | What type of assessment was used to measure the physical fitness of the participant |
| 13.Effectiveness | | Describe what results were reported by the study |
| 14. Facilitators | | The facilitators to that improve the physical fitness OR factors that support or enable the implementation of the intervention reported in the study |
| 15. Barriers | | The barriers to improve the physical fitness OR factors that inhibit the implementation of the intervention reported in the study |
| 16. Safety | | Are there safety issues that have to be bared in mind when performing the program/training |
| 17. feasibility | | To what extent is the program feasible to carry out. |

| | | |
|---|---|---|
| 18. psychosocial factors | | What is the influence of the program on the psychosocial factors of the participants |

Data will be extracted from the included studies by two assessors independently, using a pre-piloted data form (3-6 studies to check consistency).

In case of missing data or incomplete studies the first assessor will contact the authors by email.

---

#### Appendix I

| |
|---|
| **Database 2 name: <u>Embase (embase.com. NB copy/paste search in Advanced. Clear page selections)</u>** |
| **Search strategy:**<br><br>('amputation'/exp OR 'amputee'/exp OR 'limb prosthesis'/exp OR (amput* OR "limb loss" OR "leg prosthes*" OR "artificial leg"):ab,ti)<br>AND<br>('lower limb'/exp OR 'leg prosthesis'/exp OR 'below knee amputation'/exp OR 'leg amputation'/de OR 'ankle prosthesis'/exp OR 'tibial knee prosthesis'/exp OR 'tibia'/exp OR ("lower extremit*" OR "lower limb*" OR leg OR legs OR ‘one-legged’ OR "below the knee" OR "below knee" OR transtibial OR ‘trans-tibial’ OR transfemoral OR ‘trans-femoral’ OR Syme OR tibia*):ab,ti)<br>AND<br>('exercise'/exp OR 'physiotherapy'/exp OR 'kinesiotherapy'/exp OR 'exercise test'/exp OR 'rehabilitation'/lnk OR (exercis* OR training OR "prosthetic rehabilitation" OR *‘rehabilitation program*’ OR ‘rehabilitation therap*’ OR ‘rehabilitation intervention*’*):ab,ti)<br>AND<br>('endurance'/exp OR 'endurance training'/exp OR 'fitness'/exp OR 'heart rate'/exp OR 'oxygen consumption'/exp OR 'aerobic capacity'/exp OR (oxygen OR Vo2 OR "metabolic cost*" OR endurance* OR stamina OR cardio* OR aerobic OR cardiac OR exertion* OR "physical performance" OR exhaustion* OR "heart rate" OR "energy expenditure"):ab,ti)) |
| **Search result (number of items retrieved): 537**<br><br>**Search date: 10-05-2021** |

| |
|---|
| **Database 3 name: <u>CINAHL (EBSCO)</u>** |
| **Search strategy:**<br><br>((MH "Amputation+") OR (MH "Amputees") OR (MH "Limb Prosthesis") OR TI (amput* OR |

| |
|---|
| "limb loss" OR "leg prosthes*" OR "artificial leg") OR AB (amput* OR "limb loss" OR "leg prosthes*" OR "artificial leg"))<br>AND<br>((MH "Lower Extremity+") OR (MH "Below-Knee Amputation") OR TI ("lower extremit*" OR "lower limb*" OR leg OR legs OR 'one-legged' OR "below the knee" OR "below knee" OR transtibial OR 'trans-tibial' OR transfemoral OR 'trans-femoral' OR Syme OR tibia*) OR AB ("lower extremit*" OR "lower limb*" OR leg OR legs OR 'one-legged' OR "below the knee" OR "below knee" OR transtibial OR 'trans-tibial' OR transfemoral OR 'trans-femoral' OR Syme OR tibia*))<br>AND<br>((MH "Exercise+") OR (MH "Physical Therapy") OR (MH "Gait Training+") OR (MH "Therapeutic Exercise+") OR (MH "Aerobic Exercises+") OR (MH "Core Exercises") OR (MH "Group Exercise") OR (MH "Lower Extremity Exercises") OR (MH "Rehabilitation") OR (MH "Exercise Test+") OR (MH "Heart Rate") OR TI (exercis* OR training OR "prosthetic rehabilitation" OR *'rehabilitation program*' OR 'rehabilitation therap*' OR 'rehabilitation intervention*'*) OR AB (exercis* OR training OR "prosthetic rehabilitation" OR *'rehabilitation program*' OR 'rehabilitation therap*' OR 'rehabilitation intervention*'*))<br>AND<br>((MH "Physical Endurance+") OR (MH "Exertion+") OR (MH "Energy Metabolism+") OR (MH "Oxygen Consumption+") OR (MH "Physical Fitness+") OR (MH "Endurance Training") OR (MH "Heart Rate")<br>OR TI (oxygen OR Vo2 OR "metabolic cost*" OR endurance* OR stamina OR cardio* OR aerobic OR cardiac OR exertion* OR "physical performance" OR exhaustion* OR "heart rate" OR "energy expenditure") OR AB (oxygen OR Vo2 OR "metabolic cost*" OR endurance* OR stamina OR cardio* OR aerobic OR cardiac OR exertion* OR "physical performance" OR exhaustion* OR "heart rate" OR "energy expenditure")) |
| **Search result (number of items retrieved): 352**<br><br>**Search date: 10-05-2021** |

| **Database 4 name:** <u>**Web of Science (copy/paste search in Advanced)**</u> |
|---|
| **Search strategy:**<br><br>TS=(amputat* OR "artificial limbs" OR "limb loss" OR "leg prosthes*" OR "artificial leg")<br>AND<br>TS=("lower extremit*" OR "lower limb*" OR leg OR legs OR "one-legged" OR "below the knee" OR "below knee" OR transtibial OR "trans-tibial" OR transfemoral OR "trans-femoral" OR Syme OR Tibia OR tibia*)<br>AND<br>TS=(Exercis* OR "Physical Therap*" OR "Endurance Training" OR training OR "prosthetic rehabilitation" OR "rehabilitation program*" OR "rehabilitation therap*" OR "rehabilitation intervention" OR "physical activity") |

| |
|---|
| AND<br>TS=( Fitness OR "Heart Rate" OR "Energy Metabolism" OR oxygen OR Vo2 OR "metabolic cost*" OR endurance* OR stamina OR cardio* OR aerobic OR cardiac OR exertion* OR "physical performance" OR exhaustion* OR "heart rate" OR "energy expenditure") |
| **Search result (number of items retrieved): 275 titles**<br>**Search date: 10-05-2021** |

| **Database 5 name: <u>Cochrane library</u>** |
|---|
| **Search strategy:**<br>("artificial limbs" OR amput* OR “limb loss” OR “leg prosthes*” OR “artificial leg”)<br>AND<br>(“lower extremit*” OR “lower limb*” OR leg OR legs OR “one-legged” OR “below the knee” OR “below knee” OR transtibial OR “trans-tibial” OR transfemoral OR “trans-femoral” OR Syme OR tibia*)<br>AND<br>(exercis* OR training OR “*prosthetic rehabilitation” OR “rehabilitation program*” OR “rehabilitation intervention*” OR “physical therap*”)*<br>AND<br>("Energy Metabolism" OR oxygen OR Vo2 OR “metabolic cost*” OR endurance* OR stamina OR cardio* OR aerobic OR cardiac OR exertion* OR “physical performance” OR exhaustion* OR “heart rate” OR “energy expenditure” OR “physical fitness” |
| **Search result (number of items retrieved): 74 titles**<br>**Search date: 10-05-2021** |
